# Dermal trypanosomes in suspected and confirmed cases of *gambiense* Human African Trypanosomiasis

**DOI:** 10.1101/2020.02.24.20026211

**Authors:** Mariame Camara, Alseny M’mah Soumah, Hamidou Ilbouldo, Christelle Travaillé, Caroline Clucas, Anneli Cooper, Nono-Raymond Kuispond Swar, Oumou Camara, Ibrahim Sadissou, Estefania Calvo Alvarez, Aline Crouzols, Jean-Mathieu Bart, Vincent Jamonneau, Mamadou Camara, Annette MacLeod, Bruno Bucheton, Brice Rotureau

**Author notes:** Corresponding author: Brice Rotureau, PhD, Trypanosome Transmission Group, Trypanosome Cell Biology Unit, INSERM U1201 & Department of Parasites and Insect Vectors, Institut Pasteur, 25 rue du Docteur Roux, 75015 Paris, France. Joint last authors (Annette MacLeod, Bruno Bucheton, Brice Rotureau).

## Abstract

**Background:** The diagnosis of Human African Trypanosomiasis (HAT) typically involves two steps: a serological screen, followed by the detection of living trypanosome parasites in the blood or lymph node aspirate. Live parasites can, however, remain undetected in some seropositive individuals, who we hypothesize are infected with *Trypanosoma brucei gambiense* parasites in their extravascular dermis.

**Methods and findings:** To test this hypothesis, we conducted a prospective observational cohort study in the *gambiense* HAT (gHAT) focus of Forecariah, in the Republic of Guinea. 5,417 subjects in this disease foci underwent serological screening for gHAT. Of these individuals, 66 were enrolled into our study, of whom 40 were seronegative, 8 were seropositive but unconfirmed, and 18 confirmed gHAT cases. Enrolled individuals underwent a dermatological examination, and had blood samples and skin biopsies taken and examined for trypanosomes by molecular and immuno-histological methods. In confirmed cases, dermatological symptoms were significantly more frequent, relative to seronegative controls. *T. b. gambiense* parasites were present in the blood of all confirmed cases but not in unconfirmed seropositive individuals. However, trypanosomes were detected in the dermis of all unconfirmed seropositive individuals and confirmed cases. After 6 and 20 months of treatment, dermal trypanosome numbers in skin biopsies of confirmed cases progressively reduced.

**Conclusions:** Our results thus highlight the skin as a potential reservoir for trypanosomes, with implications for our understanding of this disease’s epidemiology in the context of its planned elimination and highlighting the skin as a novel target for gHAT diagnostics.

## Introduction

*Trypanosoma brucei gambiense* is a protist parasite that is transmitted by the bite of the tsetse fly and causes Human African Trypanosomiasis (gHAT, also known as sleeping sickness), in Western and Central Africa [1]. The epidemiological importance of animal reservoirs of trypanosomes is not well-characterized [2] and these parasites are considered to mostly circulate in human populations in discrete endemic foci, with approximately 13 million people at risk [3]. The number of new cases has never been so low in the known epidemiological history of the disease, with only ∼1,500 new cases reported in 2017 [4], and the World Health Organization (WHO) has targeted gHAT elimination by 2030 [3]. This objective has been encouraged by the success of active surveillance efforts that relies on a two-step diagnosis: an initial serological screen, followed by microscope observation of blood, lymph or cerebrospinal fluid (CSF) to detect extracellular trypanosomes and to confirm the serological diagnosis. However, some seropositive individuals remain without a confirmed parasitological diagnosis for years. Such, individuals have been recently described as being latent cases, raising the question as to whether reservoirs of live parasites persist in these individuals [5]. The role of skin in the transmission of arthropod-borne protozoan parasites has been overlooked for several decades [6]. *T. brucei s. l*. parasites are found in the extravascular compartment of various tissues of their mammalian hosts, including the skin, albeit mostly under experimental conditions in animal models rather than during the natural progression of the disease [7]. Recently, the extravascular tropism of African trypanosomes was re-visited in light of the new molecular and imaging technologies that are now available in animal models [8-10]. Such studies have revealed that substantial quantities of trypanosomes persist within the extravascular dermis following experimental infection in mice with *T. b. gambiense* or *T. b. brucei*. These parasites can be transmitted to the tsetse vector, even in the absence of detectable parasites in the host’s blood [9]. This study also reported a retrospective screening of archived skin biopsies from a gHAT endemic region, which revealed the presence of some extravascular skin-dwelling trypanosomes [9]. However, the species of these parasites was not identified and no clinical records were available for the screened samples.

These observations raise the question as to whether *T. b. gambiense* might be found in the skin of confirmed gHAT cases, as well as in unconfirmed seropositive individuals, in regions of active disease transmission. To address this question, we performed a prospective observational study in the Forecariah district in the Republic of Guinea, which is one of the most active gHAT foci in Western Africa.

## Methods

### Ethical approval

All investigations were conducted in accordance with the Declaration of Helsinki and fulfil the STROBE criteria. Approval for this study was obtained from the consultative committee for deontology and ethics at the Institut de Recherche pour le Développement and from the National Ethical Committee of the Republic of Guinea (Study Diag-Cut-THA 032/CNERS/17 and amendment 038/CNERS/19).

### Study enrolment, screening, and case definitions

All subjects enrolled in this study came from 43 villages in the active gHAT focus of the Forecariah District, which is located in a coastal mangrove area of the Republic of Guinea and in the HAT National Control Programme Centre of Forecariah [11]. All enrolled subjects were screened from May 2017 to February 2019 in medical surveys performed by the HAT National Control Programme, according to WHO recommendations and as described previously [12]. A total of 5,417 individuals were screened with the card agglutination test for trypanosomiasis using whole blood samples (CATTwb) (Table 1). For those individuals who tested positive in the CATTwb screening test, 5ml of blood was collected in heparinized tubes and a two-fold plasma dilution series was used to determine the CATT plasma (CATTp) end titre. All individuals with CATTp end titres of 1/4 or higher then underwent a microscopic examination of lymph node aspirate, whenever cervical swollen lymph nodes were present. Blood samples of CATTp-positive individuals were also centrifuged to obtain the buffy coat layer, which was tested for trypanosomes using the mini-anion exchange centrifugation test (mAECT BC) [13]. If trypanosomes were detected using this test, the infected individual underwent a lumbar puncture and their disease stage was determined by searching for trypanosomes using the modified simple centrifugation technique for CSF and by white blood cell (WBC) counts [14]. *Gambiense* HAT patients were classified as being stage 1 (0-5 WBC/µl and absence of trypanosomes in CSF) or stage 2 (>5 WBC/µl and/or presence of trypanosomes in CSF) and were treated accordingly by the National Control Programme. Treatment consisted of Pentamidine for stage 1 patients (intramuscular injection of 4 mg/kg once daily for 7 days in adults) or Nifurtimox-Eflornithine Combination Therapy (NECT) for stage 2 patients (oral Nifurtimox at 15 mg/kg per day in three doses for 10 days and intravenous Eflornithine (α-difluoromethylornithine or DFMO) at 400 mg/kg per day in two 2h-infusions for 7 days in adults). All parasitologically confirmed cases in this study were diagnosed and treated according to WHO recommendations.

**Table 1.**
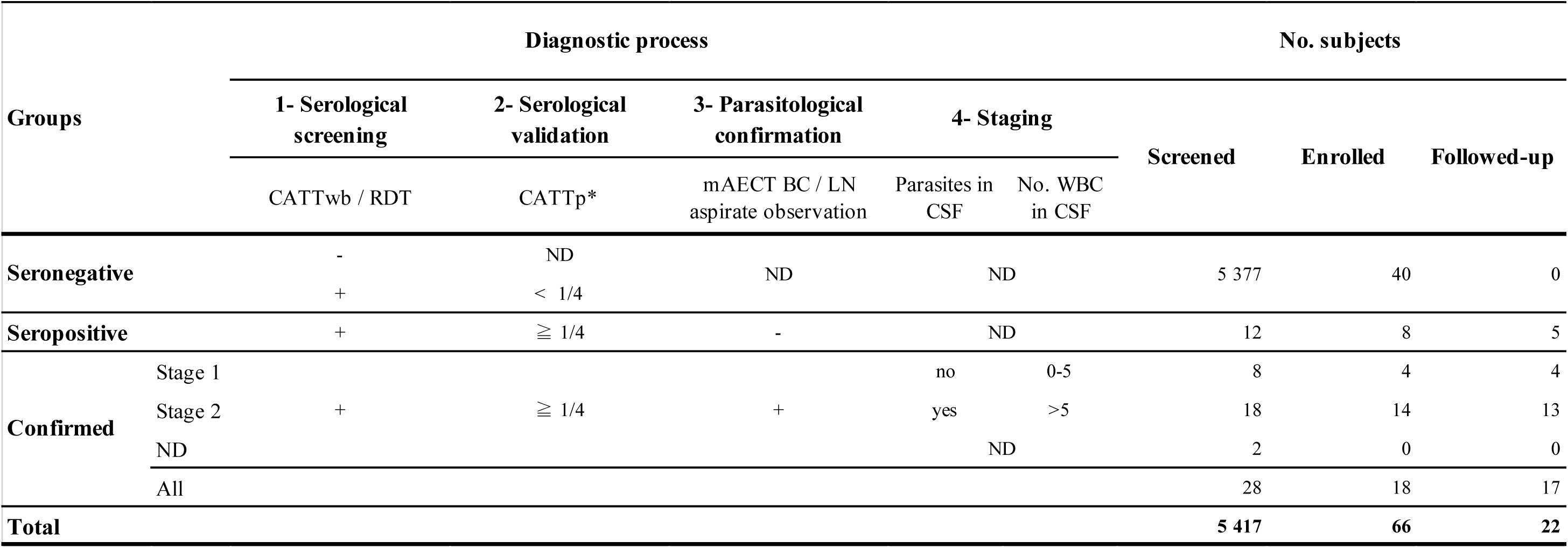
Diagnostic process, number of subjects and results. CATTwb / CATTp: card agglutination test for trypanosomiasis on whole blood / plasma; mAECT BC / LN aspirate: mini anion-exchange column technique on buffy coat / lymph node aspirate; WBC: white blood cells; CSF: cerebrospinal fluid; ND: not determined. *Highest plasma dilution with a positive result.

All confirmed cases (CATTp ≥1/4 with parasitological confirmation) and all unconfirmed seropositive individuals (CATTp ≥1/4 without parasitological confirmation) were proposed for study enrolment. Seronegative controls were randomly selected, and approximately two age-matched seronegative controls for every seropositive case were enrolled from the same village. Among the 40 seronegative controls, the first 29 individuals enrolled in the study in 2017 were only included in the epidemiological and clinical analysis but were not sampled for biological analyses, whereas the last 11 individuals enrolled in 2019 were subjected to the entire protocol, according to the recommendations from the national ethical committee of the Republic of Guinea. In total, 5/8 and 4/8 unconfirmed seropositive individuals were followed-up at 6 months and 20 months after enrolment, respectively, and 17/18 and 12/18 confirmed cases were followed-up at 6 months and 20 months after treatment, respectively (Table 4). Children under 16 years of age and pregnant women were excluded from the study. Each participant was informed about the study’s objectives in their own language and provided written informed consent. For participants between 16 to 18 years of age, informed consent was also obtained from their parents. The data of individual study participants were randomly anonymized with a 4-digit code at enrolment.

### Field procedure and sampling

At enrolment, each participant underwent an epidemiological interview and a clinical examination, during which dermatological symptoms were assessed by a trained dermatologist (AMS), as well as at each subsequent follow-up at 6 and 20 months after enrolment/treatment. This interview and clinical examination were performed for all confirmed seropositive cases, all unconfirmed seropositive individuals, and for all seronegative controls. The following epidemiological data were collected: age (in years), sex, clinical history of HAT infections in the family since 2010, and occupational risk (occurrence of any regular activities such as farming, hunting, fishing, wood-cutting and/or mining, during which an individual might be more exposed to tsetse bites). The following general clinical data were also collected during the interview: body temperature, presence of swollen cervical lymph nodes, weight loss, asthenia, eating disorders, sexual dysfunctions, repetitive headaches, circadian rhythm disruptions and/or any other behavioural changes during the last three months. Dermatological signs of pruritus (skin itch) and dermatitis (skin inflammation) were also investigated and a careful examination of the entire body was performed, in order to detect any symptoms that might be related to skin infections.

Finally, a 2mm blood-free skin punch biopsy was sampled in sterile conditions from the right back shoulder of all confirmed seropositive cases, all unconfirmed seropositive individuals, and for the final 11 seronegative controls. Biopsies were performed under local anaesthesia, and were rapidly dressed. Touch preparations were obtained by gently rolling the biopsy on a clean glass slide. The slides were air-dried, fixed in methyl alcohol, and stained with Giemsa (RAL 555 kit) for microscopic observation in the field. Biopsies were then rapidly fixed in 10% neutral buffered formalin for immuno-histochemistry and molecular analyses. Plasma aliquots from blood samples were also obtained during the screening step for use in serological trypanolysis tests, as described below.

### Immune trypanolysis test

A plasma sample from all confirmed seropositive cases, all unconfirmed seropositive individuals and from the final 11 seronegative controls was used to perform the immune trypanolysis test. This test detects complement-mediated immune responses activated by either the LiTat 1.3, LiTat 1.5 or LiTat 1.6 variable surface antigens specific for *T. b. gambiense*, as previously described [15].

### Immunohistochemical detection of trypanosomes

Skin biopsy samples were fixed in formalin, trimmed and processed into paraffin blocks in the lab. Longitudinal sections of ∼2.5μm were then prepared and processed using Dako Autostainer Plus (Dako, Denmark). Sections were stained with haematoxylin-eosin (HE) and Giemsa stains. They were also immunolabelled with the *T. brucei*-specific, anti-ISG65 antibody, which targets the Invariant Surface Glycoprotein 65 (rabbit 1/800; gift from M. Carrington, Cambridge, UK) [16], and with the *T. brucei*-specific, anti-Hsp70 antibody, which recognizes the endoplasmic reticulum molecular chaperone heat-shock protein 70 homologue (rabbit 1/2000; gift from J.D. Bangs, Buffalo, USA) [17]. For immunolabelling, the appropriate horseradish peroxidase-coupled secondary antibodies were used, and the staining was revealed with 3,3’-diaminobenzidine and counterstained with Gill’s haematoxylin. A non-infected West African skin specimen (Tissue Solutions Ltd., UK) and a *T. b. gambiense*-infected mouse skin specimen were also included with the samples as technical negative and positive controls, respectively. Immunostaining images were acquired using an Axio Observer Z1 microscope (Carl Zeiss, Germany) or a Leica 4000B microscope (Leica, Germany), and were analysed using the ImageJ 1.49v software (NIH, USA). In total, all biopsies were analysed in parallel using up to four distinct immuno-histochemical staining. The slides from each biopsy were assessed in a blinded protocol by at least two readers, with results confirmed by up to two more independent trained readers (providing an average of 8 distinct slide reads per subject at enrolment and 4 distinct reads per subject for each follow-up session). Slides from the 11 seronegative controls were randomly mixed with slides from the seropositive cases in order to guarantee blind reading. The positivity of a given skin-section slide was defined by the detection of at least three, clearly distinguishable trypanosomes.

### DNA extraction

Three, 10µm-thick sections were cut from each remaining paraffin-embedded skin specimen, with the blade being changed between samples. A non-infected West African skin specimen (Tissue Solutions Ltd., UK) and a *T. b. gambiense*-infected mouse skin specimen were also included with the samples as negative and positive technical controls, respectively. DNA was then extracted from the sections using Deparaffinization Solution (Qiagen, Germany) and the QIAamp DNA FFPE Tissue Kit (Qiagen, Germany), following the manufacturer’s instructions. For blood samples, DNA extraction was performed on 1ml blood aliquots using the DNeasy Tissue kit (Qiagen, Germany), following the manufacturer’s recommendations [18].

### PCR detection of trypanosome DNA

PCR detection of *T. brucei s. l*. parasites was performed using published primers, which hybridize to a 177bp DNA satellite repeat sequence (10,000 copies per cell) to generate a 117bp amplicon as previously described [19]. All PCR results were confirmed using new primers (TBRN3-F 5’-TAAATGGTTCTTATACGAATGA-3’ and TBRN3-R 5’-TTGCACACATTAAACACTAAAGAACA-3’) that externally flank the first primer pair to generate a larger fragment of 168bp. *T. b. gambiense*-specific PCR directed against the single copy *TgsGP* gene was also performed using published primers (308bp amplicon), as previously described [20], or by using a combination of new (5’-TATGCCGGCTACGGCACCAA-3’) and published (5’-GGGCTCCTGCCTCAATTGCTGCA-3’) [21] primers in order to reduce the amplicon to 97bp.

### Data analyses

General descriptive analyses of anonymized data were performed using Excel 16.16.11 (Microsoft, USA). Statistical analyses were performed using XLSTAT Biomed 2019.2.1 (Addinsoft, France) and Prism V.8.3.0 (GraphPad, USA) software. For epidemiological, clinical and diagnostic parameters, differences between seronegative controls versus unconfirmed seropositive individuals and confirmed cases were assessed using the following two-sided tests at 5% confidence: Fisher’s exact tests for qualitative data (Tables 2 and 3) and/or Mann-Whitney tests for quantitative data (age in Table 2). For the follow-up analyses, differences between results at enrolment versus results at 6 months and 20 months after treatment/enrolment were assessed for each group using two-sided Fisher’s exact tests at 5% confidence (Table 4).

**Table 2.**
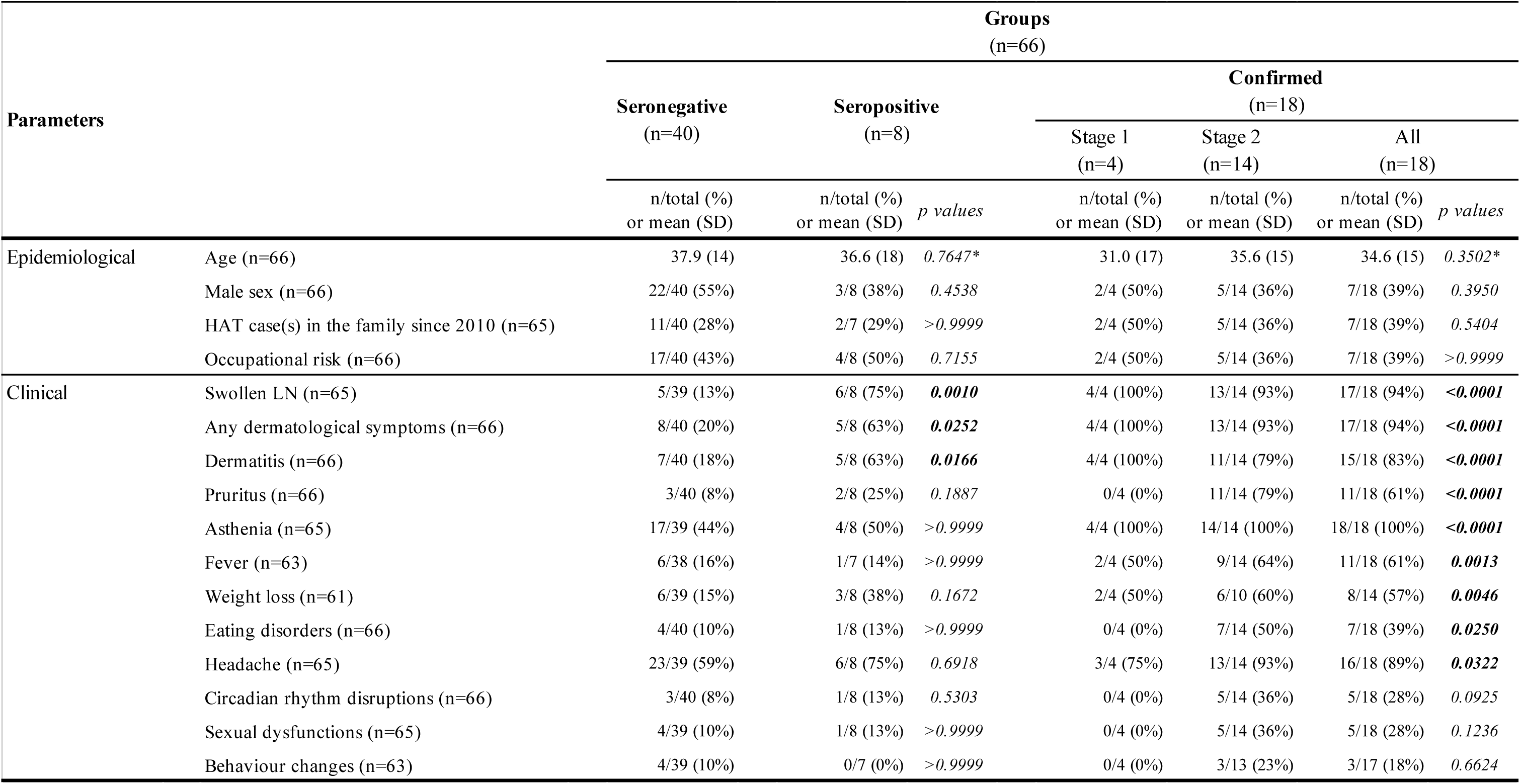
Epidemiological and clinical characteristics of case subjects. For each group and each parameter, total values correspond to the numbers of subjects for which a value was available (n/total). p values were obtained by comparing one by one the parameters of each group of seropositive subjects (unconfirmed and all confirmed) to those of seronegative controls using two-sided Fisher’s exact tests or * two-sided Mann-Whitney tests at 5% confidence. LN: lymph nodes.

**Table 3.**
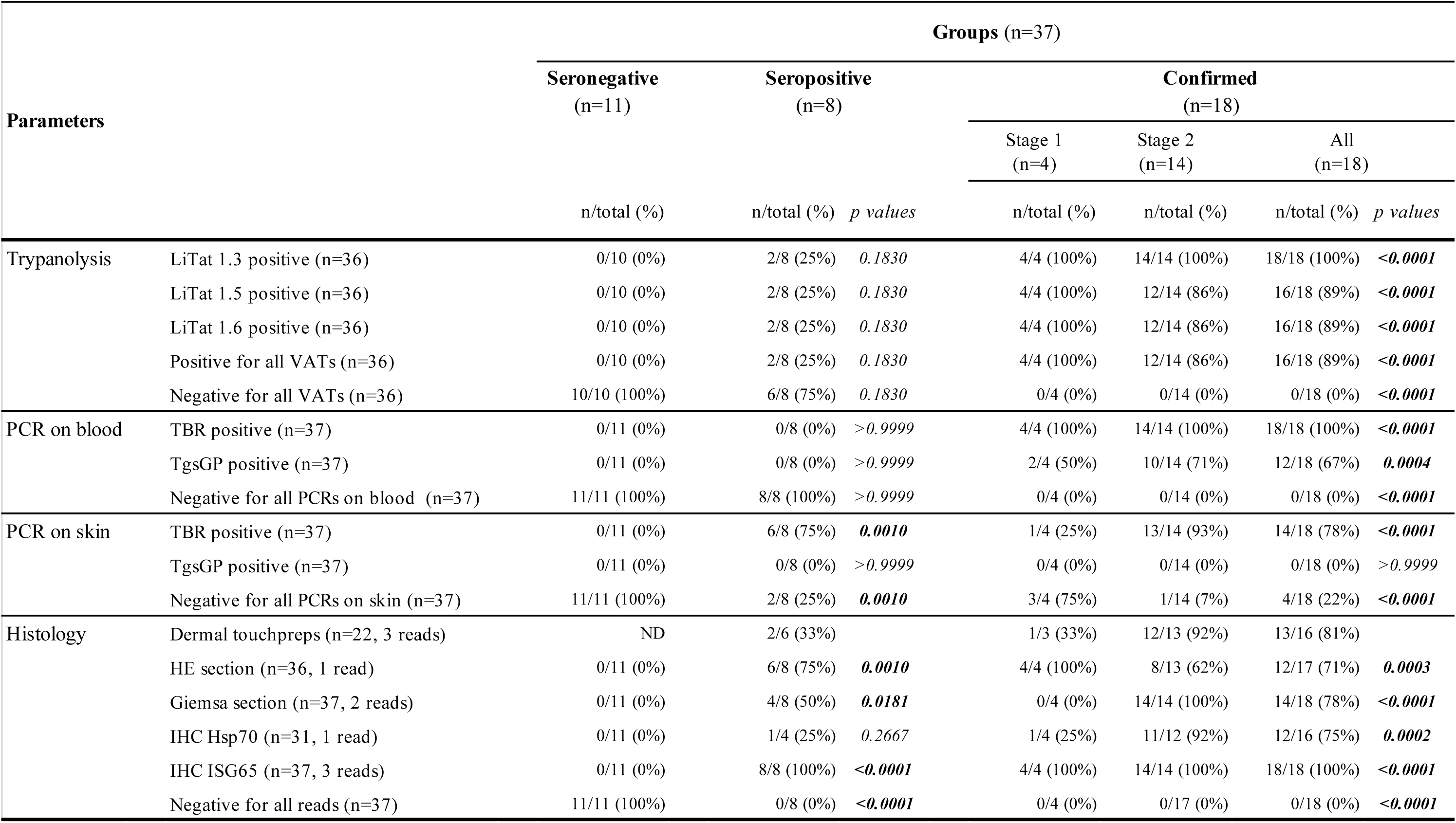
Serological, molecular and histological analysis results from blood and skin samples. For each group and each parameter, total values correspond to the numbers of subjects for which a value was available (n/total). p values were obtained by comparing one by one the parameters of each group of seropositive subjects (unconfirmed and all confirmed) to those of seronegative controls using two-sided Fisher’s exact tests at 5% confidence. VAT: variable antigen type; PCR: polymerase chain reaction; TgsGP: *Trypanosoma brucei gambiense* surface glycoprotein; HE: haematoxylin-eosin; IHC: immuno-histochemistry; Hsp70: heat shock protein 70; ISG65: invariant surface glycoprotein 65; ND: not determined.

**Table 4.**
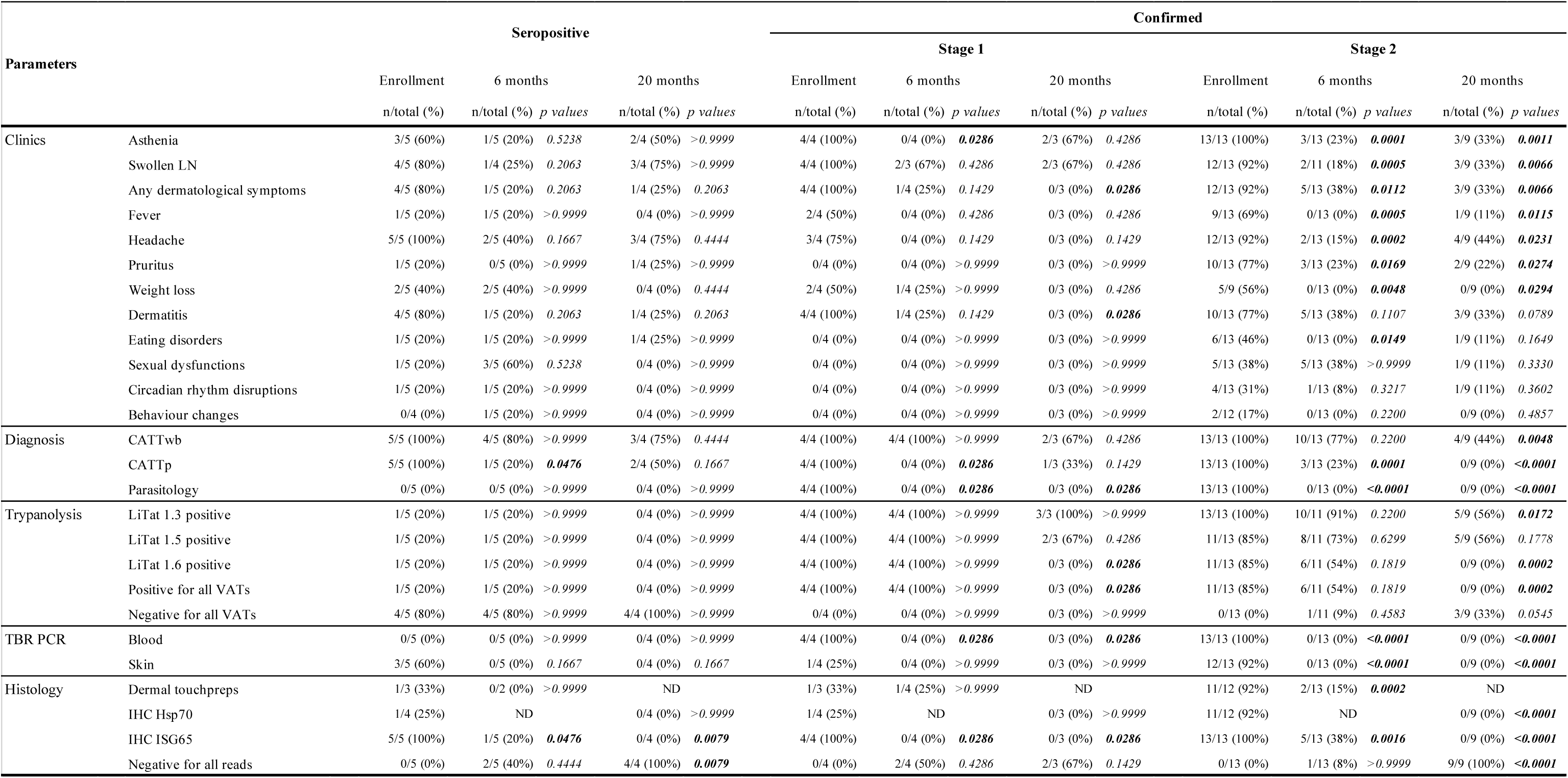
Clinical, serological, molecular and histological follow-up analyses at 6 and 20 months after enrolment. Total values correspond to the numbers of subjects for which a value was available (n/total). For each group of subjects, p values were obtained by comparing one by one the parameters recorded at 6 months and 20 months after treatment/enrolment to those obtained at enrolment, using two-sided Fisher’s exact tests at 5% confidence. LN: lymph nodes; CATT: card agglutination test for trypanosomiasis; VAT: variable antigen type; PCR: TBR polymerase chain reaction; Hsp70: heat shock protein 70; ISG65: invariant surface glycoprotein 65; ND: not determined.

## Results

### Epidemiological and clinical results

5,417 individuals were initially screened using the card agglutination test for trypanosomiasis on whole-blood (CATTwb). For CATTwb-positive individuals, a further CATT test was performed on plasma (CATTp) for serological validation. These tests identified 5,377 seronegative individuals (CATTwb-negative or CATTp<1/4), of whom 40 were enrolled as seronegative controls, of whom 11 provided skin biopsies (Table 1). Among seropositive individuals (CATTwb-positive and CATTp ≥1/4), 12 were negative for infection based on parasitological examination (prevalence of 0.22%), 8 of whom were enrolled as non-confirmed seropositive individuals (Table 1). 28 cases were found to be seropositive (CATTwb-positive and CATTp≥1/4) and further confirmed by parasitological observations (prevalence of 0.52%), including 8 stage-1 and 18 stage-2 cases, of whom 4 stage-1 and 14 stage-2 cases were enrolled as confirmed gHAT cases (Table 1). In total, 66 seronegative and seropositive (confirmed and non-confirmed) individuals were enrolled in this study and were placed into groups, depending on their screening, validation and confirmation results (Table 1).

An epidemiological and clinical assessment was performed on all enrolled subjects, including a dermatological assessment (Table 2). None of the four studied epidemiological parameters (age, sex, occupational risk, and clinical history of gHAT in the family) differed significantly between study groups (Table 2). Clinical symptoms such as asthenia (P<0.0001), fever (P=0.0013) and weight loss (P=0.0046) were statistically (Fisher’s exact test) more frequent in confirmed cases (n=18), as compared to seronegative controls (n=40) (Table 2). A particularly marked result (P<0.0001) was the presence of swollen cervical lymph nodes in 94% (17/18) of the confirmed cases and in 75% (6/8) of the unconfirmed seropositive cases, as compared to 13% (5/39) of the seronegative controls (Table 2).

Among the various observed clinical manifestations of localized dermatitis, we unambiguously identified typical cases of intertrigo (in 4/18 confirmed cases versus 2/40 seronegative controls), pityriasis (in 3/18 versus 2/40), scabies (in 3/18 versus 1/40), dermatophytosis (in 3/18 versus 1/40), molluscum (in 3/18 versus 1/40), and ulceration (in 3/18 versus 1/40). Apart from general pruritus and intertrigo, all dermatological signs were observed in upper regions of the body, especially on the thorax and arms. Confirmed cases of trypanosomiasis (83%, 15/18) were significantly more frequently affected by any types of dermatitis than were seronegative controls (18%, 7/40) (P<0.0001 by Fisher’s exact test) (Table 2). Oedematous faces were observed in four confirmed cases, and a general pruritus was reported in 64% (9/14) of the stage-2 cases, as compared to 5% (2/40) of the seronegative controls. In total, individuals with confirmed cases of gHAT presented with general or localized pruritus significantly more frequently (61%, 11/18) than did seronegative controls (8%, 3/40) (P<0.0001 by Fisher’s exact test) (Table 2). For unconfirmed seropositive individuals, an intermediate situation was observed; these individuals had a significantly higher rate of dermatitis (63%, including 1/8 with pityriasis, 2/8 with intertrigo and 3/8 with eczema) relative to seronegative controls (18%, 7/40) (P=0.0166 by Fisher’s exact test), but lower than that of confirmed cases (83%, 15/18) (Table 2).

### Biological results

Plasma from all confirmed and unconfirmed seropositive cases, and from the final 11 seronegative controls, was assessed using the trypanolysis test, which detects complement-mediated immune responses activated by *T. b. gambiense*-specific antigens. All confirmed cases were positive for the LiTat 1.3 antigen, and 89% (16/18) of these cases were positive for both the LiTat 1.5 and 1.6 antigens (Table 3). These results agree with the serological test results, which indicate that these individuals have an active trypanosome infection, at least in the blood compartment. However, only 25% (2/8) of the unconfirmed seropositive individuals were positive for all antigens; the remainder were negative for all three variants (Table 3).

Before a single 2mm skin punch biopsy was sampled from all confirmed and unconfirmed seropositive cases and from 11/40 seronegative controls, the absence of dermatitis lesions at the skin sampling site was verified. Dermal touch preparations were then generated from the biopsy samples, Giemsa-stained and read in the field. Sub-optimal ambient conditions (31±1°C at 75±6% humidity on average) during the preparation of these slides meant that their quality was sub-optimal (S1 Fig.). However, full length trypanosomes were observed on slides from 81% (13/16) of the clinical cases and from 33% (2/6) of the unconfirmed, seropositive individuals, with these results blindly confirmed by at least two independent slide readers (Table 3 and S1 Fig.). One of the unconfirmed seropositive individuals who tested positive in this dermal test, also tested positive in the trypanolysis test.

The skin biopsy samples were then fixed with formalin and later processed for immunohistochemistry analyses (IHC) in the lab. Skin samples obtained from the seronegative controls (11/11) did not test positive for trypanosomes, neither following staining by Giemsa and haematoxylin-eosin (HE) nor for the *T. brucei*-specific antibodies used in this study (Table 3). By contrast, all unconfirmed seropositive individuals (8/8) and all confirmed cases (18/18) were found to be positive at least following staining by a *T. brucei*-specific anti-ISG65 antibody targeting the Invariant Surface Glycoprotein 65 expressed at the surface of the mammalian host stages of *T. brucei s. l*. parasites (Fig. 1, S2 Fig. and Table 3). In addition, all samples from non-confirmed seropositive individuals and confirmed cases were also found to be positive following either unspecific Giemsa staining and/or unspecific HE staining and/or labelling with a *T. brucei*-specific anti-Hsp70 antibody (Fig. 1, S2 Fig. and Table 3).

**Fig. 1.**
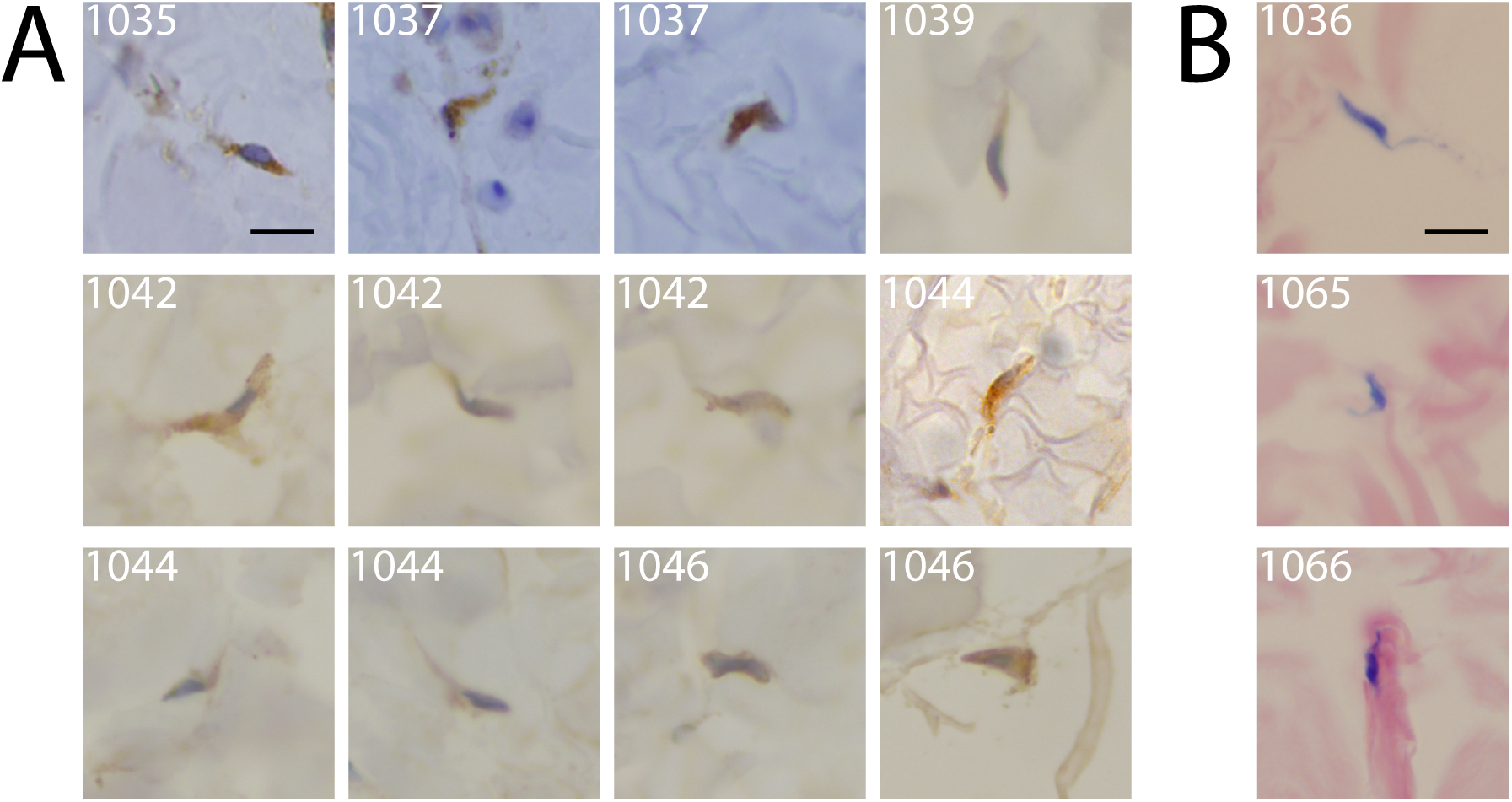
Extravascular trypanosomes in the dermal matrix of human skin biopsies. For each enrolled study subject, paraffin-embedded skin biopsy sections were stained either with (A) a specific anti-ISG65 antibody (brown) or (B) with Giemsa (purple) and screened at a 100x magnification. Representative trypanosome sections from confirmed stage-1 (subject 1044) and stage-2 cases (subjects 1035, 1036, 1037, 1039 and 1042), as well as from unconfirmed seropositive individuals (subjects 1046, 1065 and 1066) are shown. The scale bars represent 10μm. More images of extravascular *T. brucei* parasites in human skin biopsies are available in S2 Fig.

In positive skin sections, *T. brucei* parasites were evenly distributed in the reticular dermis, and were occasionally associated with oedema. To verify the size of the observed trypanosome cell sections, morphometric measurements were obtained from Giemsa-stained and ISG65-labelled skin-sections (Fig. 1 and S2 Fig.). These measurements show that the mean parasite length was 15.0 ±4.1 μm (n=293) in Giemsa-stained images and 15.9 ±3.7μm (n=109) in ISG65-labelled images.

To confirm the identity of these skin-dwelling parasites, *T. brucei*-specific PCR (TBR-PCR) assays were performed on total DNA extracted from fresh blood and from paraffin-embedded skin samples. Both blood and skin DNA samples from the seronegative controls (11/11) were found to be negative for trypanosome DNA by the TBR-PCR assays. By contrast, 100% of blood (18/18) and 78% of skin samples (14/18) from confirmed cases tested positively in the TBR-PCR assays. However, parasite DNA was only detected in the skin of unconfirmed seropositive individuals (75%, 6/8) but not in their blood (0/8) (Table 3). The less sensitive *T. b. gambiense*-specific TgsGP-PCR, which amplifies a single-copy gene, was performed on the same DNA samples and was positive for only 67% (12/18) of the blood samples of confirmed cases (Table 3). We reasoned that the use of fresh skin biopsies would be more appropriate for TgsGP-PCR. To test this hypothesis, we therefore obtained fresh skin samples, conserved in stabilization buffer, from an outgroup of nine additional confirmed cases, who were identified in 2018 using the same study protocol and who were living in the same district (S1 Table). These fresh skin samples from 89% (8/9) of these confirmed cases were found to be positive for trypanosome infection by TBR-PCR, while three cases (33%) were also found to be positive by TgsGP-PCR (S1 Table).

### Follow-up results

The same panel of analyses were repeated at 6 and 20 months after study enrolment of the unconfirmed seropositive individuals, and at 6 and 20 months after treatment of the confirmed cases (Table 4). Most of the clinical symptoms associated with the stage-2 cases at enrolment significantly decreased in frequency during the first 6 months after treatment (Table 4). A similar trend was observed for the stage-1 cases, with a statistically significant decrease in the frequency of dermatitis 20 months after treatment (P=0.0286 by Fisher’s exact test) (Table 4). The clearance of parasites from the blood and skin of all stage-2 cases who were followed-up at 20 months after treatment was assessed by a statistically significant reduction (Fisher’s exact test) in the number of positive results from the following assays: CATTwb (from 13/13 at enrolment to 4/9 at 20 months, P=0.0048), CATTp (from 13/13 to 0/9, P<0.0001), parasitological examination of body fluids (from 13/13 to 0/9, P<0.0001), trypanolysis test (from 13/13 to 5/9 LiTat1.3-positive, P=0.0172), TBR-PCR assays on blood (from 13/13 to 0/9, P<0.0001) and skin (from 12/13 to 0/9, P<0.0001), and by IHC analyses following labelling with an anti-Hsp70 antibody (from 11/12 to 0/9, P<0.0001) and an anti-ISG65 antibody (from 13/13 to 0/9, P<0.0001) (Table 4). Parasitological observations and PCR results became negative within 6 months of treatment in all confirmed cases (17/17) (Table 4). Twenty months after treatment, all confirmed cases (12/12) returned a negative result in the trypanolysis test against LiTat 1.6 and in the IHC analyses of skin sections (Table 4). However, at the end of the study, 67% (2/3) of the stage-1 cases and 44% (4/9) of the stage-2 cases still had positive results in the CATTwb test, and 100% (3/3) of the stage-1 cases and 56% (5/9) of the stage-2 cases, still had positive results in the trypanolysis test against LiTat 1.3 (Table 4). In addition, swollen lymph nodes were still detectable in 2 out of 3 stage-1 and in 3 out of 9 stage-2 cases (Table 4). For the 5 unconfirmed seropositive individuals who were monitored after enrolment, no obvious variations were observed in any of their clinical parameters over time (Table 4). Although 2 out of 4 unconfirmed seropositive individuals were found to still be positive for CATTp 20 months after enrolment, their TBR-PCR results on skin DNA and IHC staining results became negative over time. Of the six individuals who were in the unconfirmed seropositive group (n=8), and who tested positive for dermal trypanosomes by both TBR-PCR and IHC analyses at enrolment, three were lost for follow-up before 6 months (due to one death, one pregnancy and one resignation). In addition, the unconfirmed seropositive subject, who was still positive after 6 months for the CATTp, the trypanolysis test and the skin biopsy IHC with the anti-ISG65 antibody, refused to participate at the 20 months follow-up. However, during an active surveillance campaign in November 2019 (i.e. after the end of this study), this individual was diagnosed as being a stage-1 confirmed case (CATTwb +, CATTp 1/8, mAECT BC +, CSF - and WBC 4) and was treated accordingly.

## Discussion

Here, we set out to investigate whether *T. b. gambiense* parasites might be found in the skin of confirmed gHAT cases, as well as in unconfirmed seropositive individuals, in regions of active disease transmission. Although this study is somewhat limited to a restricted population and to the detection methods used, 100% of the confirmed cases and unconfirmed seropositive subjects were found to carry extravascular trypanosomes in their skin.

### Detection of trypanosomes

Routine molecular analyses by TBR-PCR confirmed the presence of *T. brucei s. l*. parasites in both the blood and skin of confirmed cases. However, parasite DNA was only detected in the skin of unconfirmed seropositive individuals, which might underlie their infectious status in the extravascular compartment. In contrast, no positive results were found by *T. b. gambiense*-specific TgsGP-PCR on DNA extracted from paraffin-embedded skin samples, likely due to the low sensitivity of this method, which targets a single-copy gene. However, the positivity of some direct TgsGP-PCR assays, performed on fresh skin DNA samples from an outgroup of confirmed cases, suggests that the trypanosomes found in the dermis of the confirmed cases enrolled in the present study are likely to be *T. b. gambiense* parasites.

The trypanolysis test results were also compatible with previous reports [12]. Nevertheless, it is noteworthy that, within the limits of the method used, 6 out of 8 unconfirmed seropositive subjects gave a negative trypanolysis test result, despite the presence of skin-dwelling parasites in these individuals, as confirmed by histological approaches. In 2 of these unconfirmed seropositive subjects who had a positive trypanolysis test result, trypanosomes were also detected in their skin samples by at least 2 distinct histological approaches and by PCR. These observations suggest that unconfirmed seropositive subjects who have negative trypanolysis test results carry trypanosomes in the extravascular compartment. In addition, although anti-LiTat1.3 antibodies were detected by CATT in the plasma of these unconfirmed seropositive subjects, the trypanolytic activity of these anti-LiTat1.3 antibodies appeared limited as assessed by the negativity of the corresponding trypanolysis tests. The apparent lack of congruence between results of the CATTp and of the trypanolysis test is possibly reflecting a peculiar status of the immune response in some non-confirmed seropositive subjects that remained to be studied more in-depth.

### Dermatological signs in gHAT

Our results indicate that dermatological symptoms might be an important aspect of gHAT’s clinical presentation. The few reports that exist on this topic in the literature describe a wide array of skin pathologies associated with sleeping sickness, including pruritus, chancre, rashes and localized oedemas [22, 23]. However, detailed dermatological profiles of HAT cases have mostly been derived from light-skinned travellers with imported HAT, who apparently experience a more rapid disease onset and more skin manifestations than do dark-skinned African patients [23]. Whereas chancres and rashes remain anecdotal, pruritus was the most commonly observed dermatological sign in endemic cases (this condition is present in up to 57% of stage-2 cases) [23]. In our study, we observed a higher occurrence of pruritus and dermatitis in unconfirmed seropositive individuals and in confirmed cases, relative to seronegative controls (Table 2). The observed dermatitis profiles included some conditions the aetiologies of which might not be directly related to a trypanosome infection. However, it could be hypothesized that the immune status of the infected host skin is somehow altered by the presence of trypanosomes in a way that promotes the outcome of dermatitis caused by other pathogens and/or increases skin sensitivity. The direct detection of trypanosomes in the human skin is not well documented in the literature [22]. In our immunohistochemical analyses of skin sections, only a limited portion of each parasite cell is visible, because entire trypanosomes do not necessarily lie in the same plan as the 2.5μm skin sections. For the same reason, the parasite nucleus, kinetoplast and flagellum are rarely all visible in the same given cell section. However, the specificity of the anti-ISG65 and anti-Hsp70 antibodies used in this study enabled the slide readers to unambiguously detect most trypanosomes within the extracellular dermal matrix in a blinded analysis of skin samples. The detection of skin-dwelling parasites at enrolment in most of the 2mm skin punch biopsies sampled from seropositive individuals indicates that skin-dwelling parasites might be present over a considerable proportion of the skin surface. In this study, the choice of the sampling site was guided by preliminary observations recorded in the clinical files of previously confirmed cases in the same region; these cases presented with pruritus more frequently in the upper regions of the body. We reasoned that these bouts of itching could have been provoked by dermal trypanosomes, hence the choice of the right shoulder as the skin sampling site in this study. However, the precise dynamics of parasite load and distribution in the extravascular dermal compartment over the course of an infection remains unknown. According to historic (reviewed in [7]) and more recent [9] studies in experimental animal models, skin-dwelling parasites could theoretically be detected in almost the entire skin surface, yet with a variable distribution and at variable local densities.

### A dermal reservoir of trypanosomes in non-confirmed seropositive individuals

One possible explanation for the persistence of disease foci in certain regions is the presence of animal reservoirs [2]. Another possibility, as increasing evidence suggests, is that traditionally used diagnostic approaches do not detect some *T. b. gambiense* infections among seropositive cases [2]. Indeed, bloodstream parasite numbers in *T. b. gambiense* infections can periodically fluctuate to less than 100 trypanosomes/ml, falling below the detection limit of the most sensitive methods currently in use [24]. Another study estimated that 20-30% of gHAT cases are missed in active case detection by standard parasitological techniques and are left untreated [25]. These infected individuals might ultimately progress to clinical disease or remain almost asymptomatic until undergoing a possible self-cure [2]. Here, for the first time, we provide parasitological evidence for the presence of trypanosomes in eight unconfirmed seropositive subjects. The presence of trypanosomes in the skin of these individuals suggests that the human reservoir of gHAT might be greater than initially thought. In this study, the negative test and assay results obtained on samples from the five unconfirmed seropositive subjects who were followed-up at 20 months suggest that these individuals might have been enrolled during a transient/ending/self-curing infection. In this context, data on the infectious status of the 3 remaining unconfirmed seropositive subjects who were found to have dermal trypanosomes at enrolment, but who were lost from follow-up, would have been informative. However, there remains the case of the unconfirmed seropositive individual, who remained positive for dermal parasites 6 months after enrolment, and who was diagnosed as a confirmed stage-1 case in an active screening campaign 31 months after enrolment, after this study had ended. This individual’s diagnosis suggests that dermal parasites could play a significant role during the early stage of the disease.

### Transmission and epidemiological contribution of dermal trypanosomes

Mathematical modelling recently predicted that, in the absence of any animal reservoirs, these unconfirmed seropositive individuals could contribute to disease transmission by maintaining an overlooked reservoir of skin-dwelling parasites [26]. Tsetse feed by lacerating the skin of their host rather than by inserting a proboscis directly into the vasculature. After causing significant local damage at the bite site, the insects ingest the resultant pool of capillary blood and lymph mixed with the surrounding dermal material. Bearing in mind this feeding mechanism and the observation that latent cases host dermal trypanosomes, the skin of such individuals could provide a population of infective parasites that are readily accessible to the tsetse fly. Indeed, this mode of transmission has been demonstrated in experimental animal models, in which skin-dwelling trypanosomes were efficiently transmitted to the tsetse vector, even in the absence of detectable parasitaemia [8, 9, 27]. However, the presence of the parasite forms (called stumpy trypanosome stages) that are assumed to be most adapted for development in tsetse flies were not investigated here. This is an important question for future studies to address, in order to estimate the actual infectivity potential of human skin-dwelling parasites. Our reported observations should also be confirmed in a larger number of unconfirmed seropositive individuals and seronegative controls, and the study scaled-up to include other endemic transmission foci in Africa, in order to confidently determine the actual prevalence of dermal trypanosomes.

### Conclusion

Our results also raise questions about the strategies used to diagnose this disease, which currently focus on detecting parasites in the blood and lymph. If the human skin is indeed a disease reservoir, it could represent a novel target for diagnostics, and it could: (i) allow more carriers to be treated; (ii) help to determine a more accurate estimate of the true prevalence of the disease; and (iii) help to identify as yet undetected reservoirs in both human and animal populations. The current WHO recommendation, based on risk-benefit analyses, is to not treat unconfirmed seropositive individuals without knowing if they have an active infection [1]. Importantly, we observed that the routinely administered trypanocide treatments (Pentamidine for stage-1 and NECT for stage-2 cases, according to the previous WHO recommendations) efficiently targeted both bloodstream and dermal trypanosomes in all the patients followed-up, as shown by the negative results of all the tests used in this study after 20 months. With the promise of new cheaper, less toxic and easier to administer drugs on the horizon, the policy of treating unconfirmed seropositive individuals could possibly be reconsidered. Indeed, the drug Acoziborole, which requires a single oral administration, could hopefully be the next revolutionary treatment against gHAT. As gHAT approaches its elimination targets, we propose from our findings that current algorithms, which are used to identify and manage disease cases, could be adapted to include the detection of skin-dwelling parasites, which likely represent a previously unaccounted for anatomical reservoir.

## Data Availability

Upon request, the original protocol and associated forms, as well as an anonymised dataset, could be obtained from the corresponding author.

## Acknowledgements

We thank M. Carrington (Cambridge, UK) and J.D. Bangs (Buffalo, USA) for providing antibodies. We are grateful to Y. Madec (Paris, France) for his help in statistical analysis. We warmly thank the team of the Programme National de Lutte contre la Trypanosomiase Humaine Africaine of Conakry, as well as all our collaborators of the Forecariah District Health Department. We thank Dominique N’Diaye (Paris, France) for his critical reading and Jane Alfred (Catalyst Editorial, UK) for her editorial work on the manuscript.

## Author contributions

MarC, AMS and NRKS conduced the clinical study in the field and commented on the manuscript. HI, IS, CT, CC, ACo, ACr, OC, ECA and JMB performed sample analyses and commented on the manuscript. MamC and VJ held logistical aspects, analysed part of the data and commented on the manuscript. AML*, BB* and BR* designed the study, organised logistical aspects, analysed the data and wrote the manuscript as co-last authors.

## Transparency statement

The lead author affirms that the manuscript is an honest, accurate and transparent account of the study being reported, that no important aspect of the study has been omitted, and that any discrepancies from the study as originally planned have been explained.

## Supporting information legends

**S1 Fig. Giemsa-stained skin biopsy touch-preparations showing trypanosomes among extracellular dermal matrix elements**. For most of the enrolled seropositive subjects, blood-free skin biopsy touch-preps were directly stained with Giemsa in the field and scrutinized at a 100x magnification. Scale bars represent 10μm and white arrows indicate dermal trypanosomes.

**S2 Fig. Pictures of skin biopsy sections with different immuno-histological staining showing extravascular *T. brucei* parasites in the dermal matrix of both unconfirmed seropositive individuals and confirmed cases**. For each enrolled subject identified by a 4-digit code, paraffin-embedded skin biopsy sections were either counterstained with Giemsa, HE, an anti-ISG65 antibody or an anti-Hsp70 antibody and scrutinized at a 100x magnification. Scale bars represent 10μm and white arrows indicate dermal trypanosome sections.

**S1 Table. Detailed serological and molecular analysis results from blood and skin samples of additional confirmed cases**. CATTwb / CATTp: card agglutination test for trypanosomiasis on whole blood / plasma; mAECT BC: mini anion-exchange column technic on buffy coat; PCR: single round polymerase chain reaction; TgsGP: *Trypanosoma brucei gambiense* surface glycoprotein; +: positive result; -: negative result.

